# Studies of Novel Coronavirus Disease 19 (COVID-19) Pandemic: A Global Analysis of Literature

**DOI:** 10.1101/2020.05.05.20092635

**Authors:** Bach Xuan Tran, Giang Hai Ha, Long Hoang Nguyen, Giang Thu Vu, Hai Thanh Phan, Huong Thi Le, Carl A. Latkin, Cyrus S.H. Ho, Roger C.M. Ho

**Affiliations:** Institute for Preventive Medicine and Public Health, Hanoi Medical University, Hanoi 100000, Vietnam; Bloomberg School of Public Health, Johns Hopkins University, Baltimore, MD 21205, United States; Institute for Global Health Innovations, Duy Tan University, Da Nang 550000, Vietnam; Faculty of Medicine, Duy Tan University, Da Nang 550000, Vietnam; VNU School of Medicine and Pharmacy, Vietnam National University, Hanoi 100000, Vietnam; Center of Excellence in Evidence-based Medicine, Nguyen Tat Thanh University, Ho Chi Minh City 700000, Vietnam; Department of Psychological Medicine, National University Hospital, Singapore 119074, Singapore; Institute for Health Innovation and Technology (iHealthtech), National University of Singapore, Singapore 117599, Singapore; Department of Psychological Medicine, Yong Loo Lin School of Medicine, National University of Singapore, Singapore 119228, Singapore

**Keywords:** Scientometrics, content analysis, text mining, COVID-19

## Abstract

An exponential growth of literature about novel coronavirus disease 19 (COVID-19) has been observed in the last few months. This textual analysis of 5,780 publications extracted from the Web of Science, Medline, and Scopus databases was performed to explore the current research focuses and propose further research agenda. The Latent Dirichlet allocation was used for topic modeling. Regression analysis was conducted to examine country variations in the research focuses. Results indicated that publications were mainly contributed by the United States, China, and European countries. Guidelines for emergency care and surgical, viral pathogenesis, and global responses in the COVID-19 pandemic were the most common topics. There was variation in the research approaches to mitigate COVID-19 problems in countries with different income and transmission levels. Findings highlighted the need for global research collaboration among high- and low/middle-income countries in the different stages of prevention and control the pandemic.

## Introduction

Novel coronavirus disease 19 (COVID-19), caused by severe acute respiratory syndrome coronavirus 2 (SARS-CoV-2), is currently threatening millions of lives in the world. After being introduced at the end of 2019, this disease was officially declared as a global pandemic by the World Health Organization (WHO) on March 11, 2020.^1^ Until April 30, 2020, 185 countries/territories reported 3·2 million confirmed cases with 227,847 total deaths,^2^ and the highest-burden has been placed in European and American countries.^1^ Serious health, social and economic consequences of COVID-19 have been well-recognized,^3-7^ especially among the elderly with comorbidities, homeless individuals, and also residents who face financial, mental, and physical hardships due to social distancing policies.^8^

Given that COVID-19 is a new threat without any antiviral therapies or vaccines, current measures to mitigate this crisis depend heavily on the national and regional preparedness and responses.^9^ However, optimal strategies to cope with the complexity of this pandemic demands substantial scientific evidence. Recently, the WHO has issued technical guidance for countries/regions and research institutions, as well as worked closely with global researchers to update the empirical evidence.^10,11^ Efforts have been made around the globe to enhancethe understanding of the dynamic transmission of COVID-19, appropriate vaccine research strategies effective treatments, as well as the impacts of current responses on population health and well-being.^12^ As a result, in the last four months, the number of publications about COVID-19 has been increased dramatically, including articles, reviews, letters to editors, or preprint documents.^13^ These contributions have proven the importance of scientific research in pandemic preparedness as well as helping the governments to respond rapidly and effectively to the crisis.^14^

Cumulatively, current research evidence has partly shaped our knowledge about COVID-19, but there has been still raising more questions to address, which require sharing information and providing scientific expertise and leadership of all countries to accelerate research efforts.^15^ Yet, there has been a the lack of studies attempted to identify the research focus in different countries and regions. Previous systematic reviews have been conducted to explore in detail the clinical characteristics of COVID-19,^16,17^ or the effectiveness of specific COVID-19 treatment,^18,19^ and policies.^20^ However, these studies were conducted on a small volume of articles. One potential approach to delineate the focus of the academic community is bibliometric analysis. This method has been previously applied to identify the rapid development of research output, the most prolific countries, journal information, and major concerns of research, but not provide a deep analysis of publications.^21-23^ In this study, by using text visualization and topic modeling approaches as a part of natural language processing and machine learning, we aimed to explore the research focus in general and in countries with different levels of income and COVID-19 transmission features. Findings might provide a knowledge gap in scientific research related to COVID-19 and future direction to inform research focus and policy responses better.

## Materials and Methods

### Searching strategy and selection criteria

Information on COVID-19 and SARS-CoV-2-related documents published until 23 April 2020 were extracted from the Medline, Scopus, and Web of Science (WoS) databases. The search terms and search queries for each online database were developed according to the WHO naming process for the virus and the disease it causes,^24^ and are presented in the **Supplementary 1**. Any English-language publications containing COVID-19 disease or SARS-CoV-2 virus published from December 2019 to 23 April 2020 were included. Document types such as corrections, data papers, reprints, or conference papers were excluded because they might be duplicated to peer-review papers. Datasets of three databases were merged, and duplications were screened independently and removed by two researchers. A final dataset of 5,780 papers was used for further analysis. The searching process was presented in **Figure 1**.

**Figure 1.**
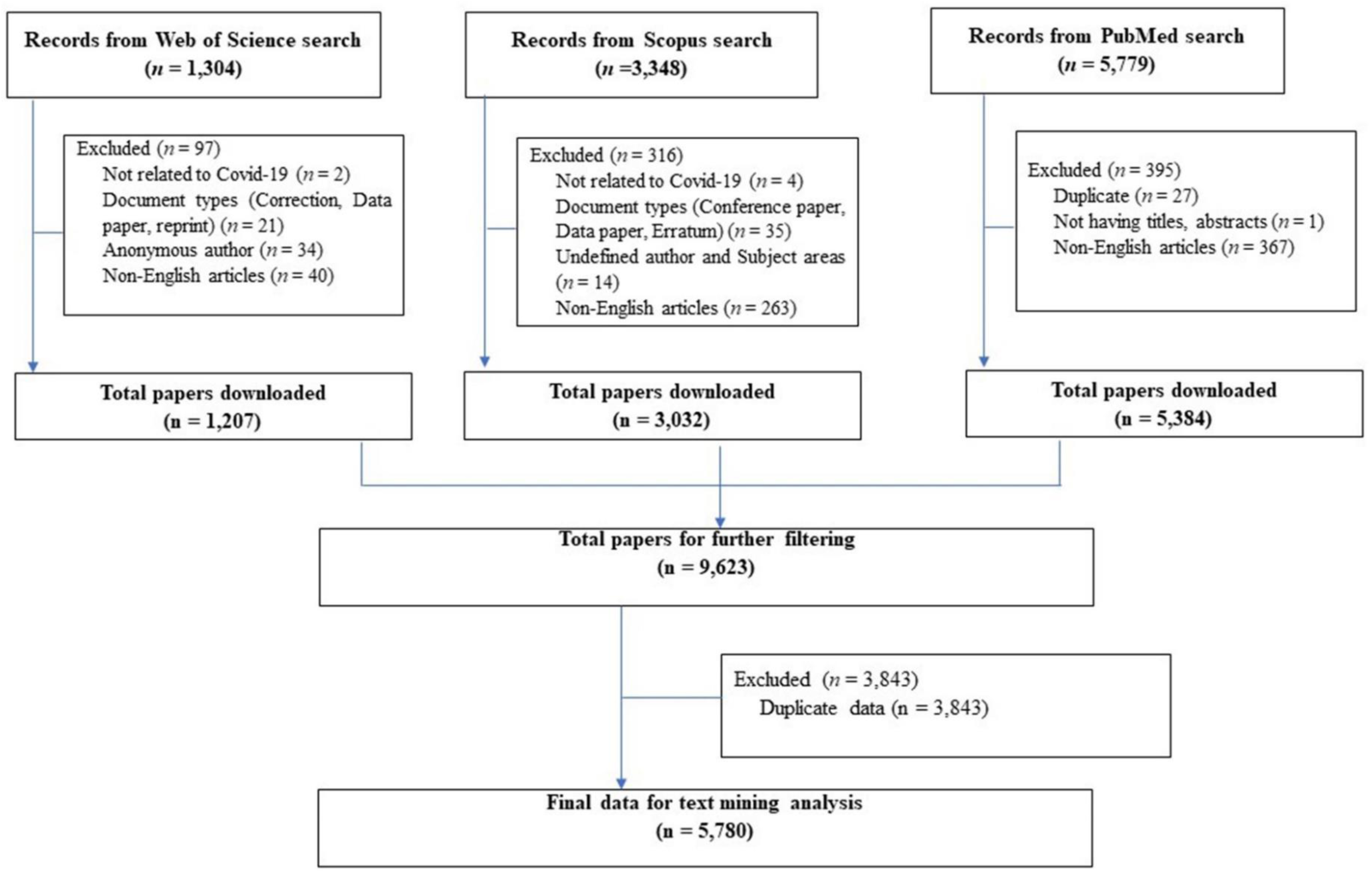
Selection process.

### Data analysis

In this paper, we extracted the documents’ title, abstract, keywords, citation, and affiliation of authors for analysis. As a document could be authored by scholars from different countries, we considered that all these countries contributed to the document preparation. Moreover, we decided to include both documents with and without abstracts for text analysis since the title of the document could partly reflect the document’s topic. We first descriptively analyzed the number of publications in each country and presented these data by using Microsoft Excel’s Map function. Then, we exported the top ten most cited publications for a detailed analysis of the content of the most cited papers.

We used the VOSviewer textual analysis software tool (version 1·6·15, Centre for Science and Technology Studies, Leiden University, the Netherlands) to measure the co-occurrence of keywords and most frequent terms in title/abstract.^25,26^ Then, we employed Latent Dirichlet allocation (LDA) to discover fifteen latent topics from the titles and abstracts of documents. This Bayesian model treats each document as a set of topics, and topics are probability distributed over a set of words and their co-occurrence.^27^ Thus, the LDA technique can produce two outputs: 1) Probability distributions of different topics per document (to acknowledge how many topics are created based on the given publications), and 2) Probability distributions of unique words per topic (to define the topics).^27^ Because each title/abstract may contain a mixture of topics, the LDA outputs may not reflect a specific research field or discipline. However, experience for previous work suggested that documents that focused on particular themes would be more likely to be categorized in the same group. To ensure robust results in naming the topic, we also checked at least ten documents per topic to ensure that the theme could fit the content of documents.

Multivariable linear regression models were performed to examine the research focus of countries with different income classification (low, low-middle, high-middle, and high income – according to World Bank classification),^28^ and different transmission classification (Pending, Sporadic case, Clusters of cases, Community transmission – according to WHO).^29^ The dependent variable was the share of publications in specific topic out of total publications in each country (%), while the independent variables were income classifications and transmission classifications. The models were adjusted to the natural logarithm of gross domestic product (GDP) per capita, the number of COVID-19 cases, and the number of COVID-19 deaths per country. The latest data on GDP per capita and income classifications were collected from the World Bank databases, while data on cases deaths were extracted from WHO reports on April 24, 2020. A p-value of less than 0.05 was used to detect statistical significance.

## Results

**Figure 2** shows the research productivity of each country. 115 countries produced 5,780 publications in the searching period. It appears that scientific publications were mainly driven by the research hubs such as China, the United States, Canada, France, Italy, the United Kingdom, and India, which were also heavily hit by COVID-19. In contrast, the majority of African countries had no more than ten studies about COVID-19.

**Figure 2.**
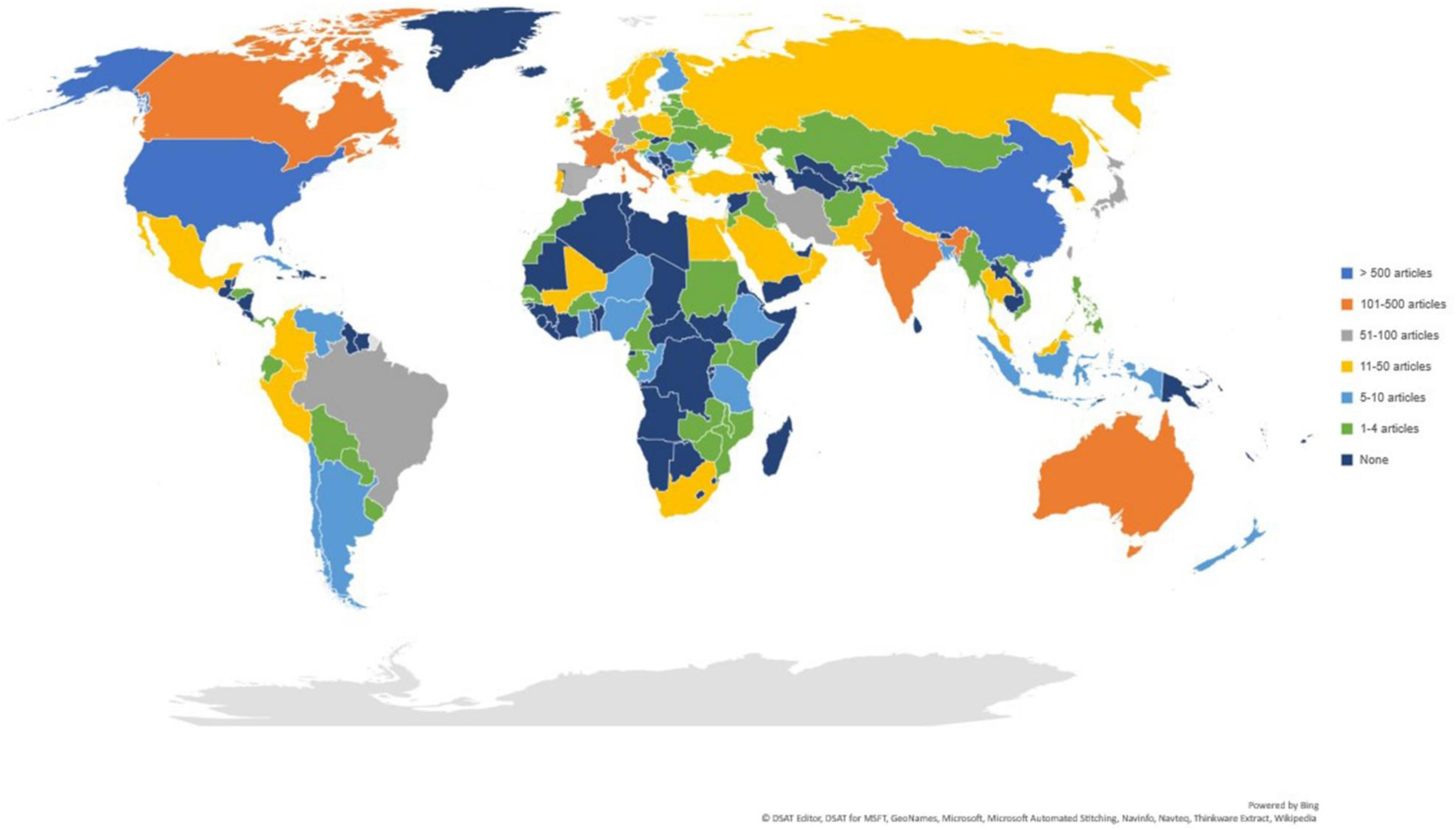
Number of publications per country.

The list of ten most cited publications about SARS-CoV-2 and COVID-19 and their main findings are presented in **Table 1**. Reports on the clinical and laboratory characteristics of the confirmed cases are of most interest, with six out of ten papers in the list. The most cited paper was a descriptive study about epidemiological and clinical features of 99 cases from Wuhan, China, which was believed to be the genesisof SARS-CoV-2.

**Table 1.** Top ten most cited papers

| No. | Title | Journal (IF) | Number of Citation | Main findings |
| --- | --- | --- | --- | --- |
| 01 | Epidemiological and clinical characteristics of 99 cases of 2019 novel coronavirus pneumonia in Wuhan, China: a descriptive study | The Lancet (IF=59.1) | 319 | • SARS-CoV-2 infection was of clustering onset, and more likely to affect older males with comorbidities.<br>• Patients had clinical manifestations of fever, cough, shortness of breath, muscle ache, confusion, headache, sore throat, rhinorrhoea, chest pain, diarrhoea, and nausea and vomiting.<br>• Imaging examination revealed bilateral pneumonia, multiple mottling and ground-glass opacity. |
| 02 | A familial cluster of pneumonia associated with the 2019 novel coronavirus indicating person-to-person transmission: a study of a family cluster | The Lancet (IF=59.1) | 245 | • Results confirmed that SARS-CoV-2 was transmitted through person-to-person contact.<br>• Older patients (aged >60 years) had more systemic symptoms, extensive radiological ground-glass lung changes, lymphopenia, thrombocytopenia, and<br>increased C-reactive protein and lactate dehydrogenase levels.<br>Phylogenetic analysis of showed that this is a novel coronavirus, which is closest to the bat severe acute respiatory syndrome (SARS)-related coronaviruses found in Chinese horseshoe bats. |
| 03 | Clinical characteristics and intrauterine vertical transmission potential of COVID-19 infection in nine pregnant women: a retrospective review of medical records | The Lancet (IF=59.1) | 75 | Clinical characteristics of COVID-19 pneumonia in pregnant women were similar to those reported for non-pregnant adult patients.<br>Fevers, cough, myalgia, sore throat and malaise were also observed.<br>No neonatal asphyxia was observed in newborn babies. |
| 04 | Detection of 2019 novel coronavirus (2019-nCoV) by real-time RT-PCR | Eurosurveillan ce (IF=7.4) | 74 | The laboratory diagnostic workflow for detection of SARS-CoV-2 was described and validated. |
| 05 | Emerging coronaviruses: Genome structure, replication, and pathogenesis | Journal of Medical Virology (IF=2.0) | 51 | • Available understanding on genome structure and replication, and functions proteins in coronaviral replication of coronaviruses (CoVs) were reviewed.<br>• SARS-CoV-2 has a typical genome structure of CoV and belongs to the cluster of betacoronaviruses, including Bat-SARS-like (SL)-ZC45, Bat-SL ZXC21, SARS-CoV, and MERS-CoV. |
| 06 | CT imaging features of 2019 novel coronavirus (2019-NCoV) | Radiology (IF=7.6) | 51 | • Typical CT findings included bilateral pulmonary parenchymal ground-glass and consolidative pulmonary opacities, sometimes with a rounded morphology and a peripheral lung distribution.<br>• Lung cavitation, discrete pulmonary nodules, pleural effusions, and lymphadenopathy were absent. |
| 07 | Presumed Asymptomatic Carrier Transmission of COVID-19 | JAMA - Journal of the American Medical | 49 | • All symptomatic patients had multifocal ground-glass opacities on chest CT, and 1 also had subsegmental areas of consolidation and fibrosis. |
|  |  | Association<br>(IF=51.3) |  | • All the symptomatic patients had increased C-reactive protein levels and reduced lymphocyte counts.<br>• The coronavirus may have been transmitted by the asymptomatic carrier. |
| 08 | Breakthrough: Chloroquine phosphate has shown apparent efficacy in treatment of COVID-19 associated pneumonia in clinical studies. | BioScience Trends<br>(IF=1.7) | 48 | • Chloroquine phosphate is superior to the control treatment in inhibiting the exacerbation of pneumonia, improving lung imaging findings, promoting a virus-negative conversion, and shortening the disease course according to the news briefing.<br>• Severe adverse reactions to chloroquine phosphate were not noted in the patients in trial. |
| 09 | Genomic characterization of the 2019 novel human-pathogenic coronavirus isolated from a | Emerging Microbes and Infections<br>(IF=6.2) | 46 | • Genome of SARS-CoV-2 has 89% nucleotide identity with bat SARS-like-CoVZXC21 and 82% with that of human SARS-CoV. |
|  | patient with atypical pneumonia after visiting Wuhan |  |  | Phylogenetic trees of their orf1a/b, Spike, Envelope, Membrane and Nucleoprotein clustered closely with those of the bat, civet and human SARS coronaviruses. |
| 10 | Incubation period of 2019 novel coronavirus (2019- nCoV) infections among travellers from Wuhan, China, 20-28 January 2020 | Eurosurveillance (IF=7.4) | 30 | The mean incubation period was estimated to be 6.4 days (95% credible interval: 5.6–7.7), ranging from 2.1 to 11.1 days (2.5th to 97.5th percentile). |

**Figure 3** presents the network of 200 keywords with a co-occurrence of at least 20 times. The keywords were assigned to three major clusters. Cluster 1 (blue) reveals some basic imaging techniques for the diagnosis of lung function impairments (tomography and thorax radiograph) in children, adolescents, and adults. Cluster 2 (red) refers to the major concerns of the world regarding COVID-19, such as prevention, medicine, and public health response. Cluster 3 (green) focuses on the biology of SARS-CoV-2, including the origin, the phylogenetic network, and the genomic, proteomic, and metabolomic characterization of the virus.

**Figure 3.**
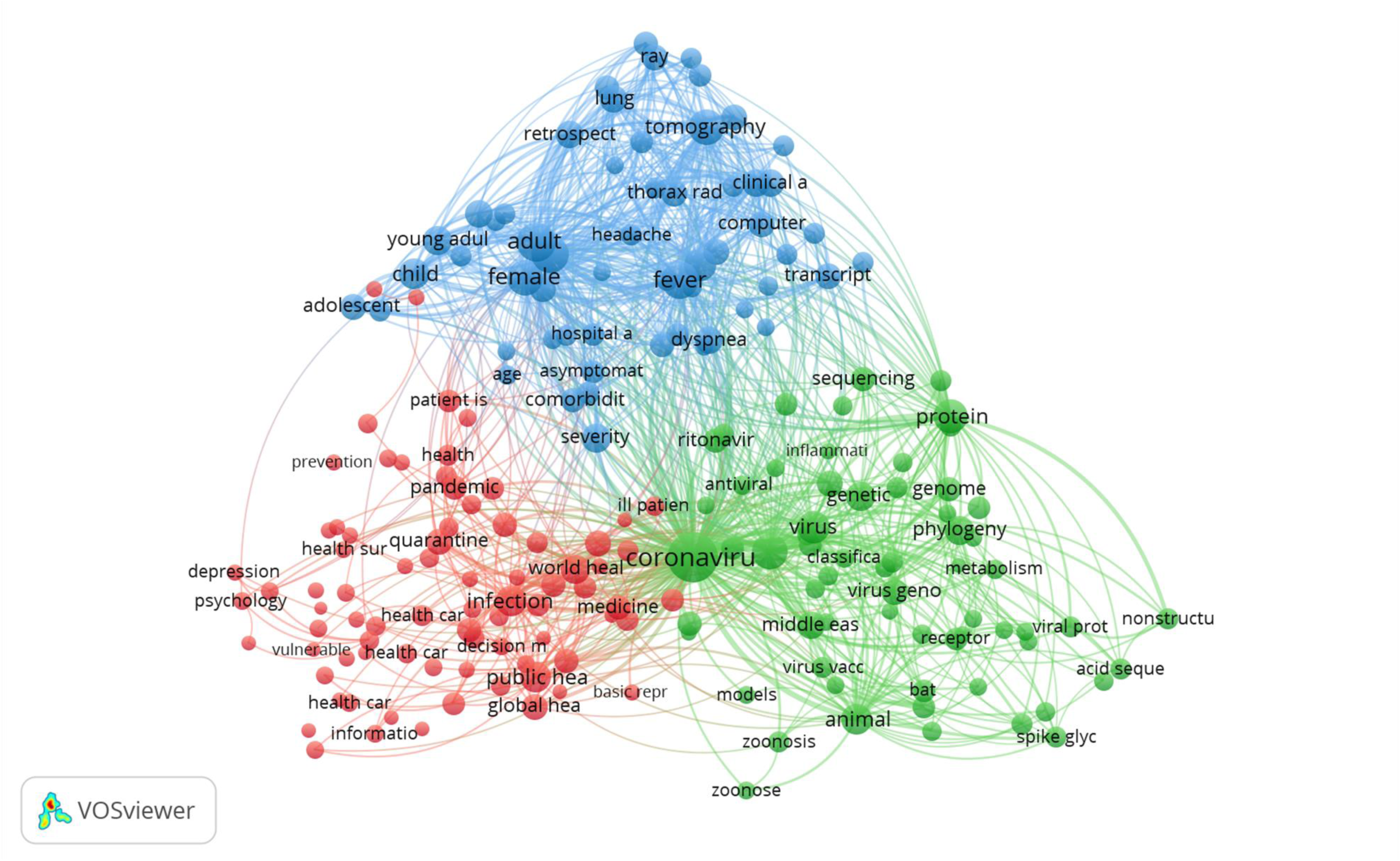
Co-occurrence analysis of keywords.

Thematic analysis of 250 most frequent terms is presented in **Figure 4**. Major themes of current research on COVID-19 are (1) promising therapies for COVID-19 prevention and treatment, and their mechanisms (blue), (2) hot spots of the pandemic and the governments’ responses (red), and (3) clinical patterns, such as ground-glass opacities (GGO), and complications of COVID-19 (green).

**Figure 4.**
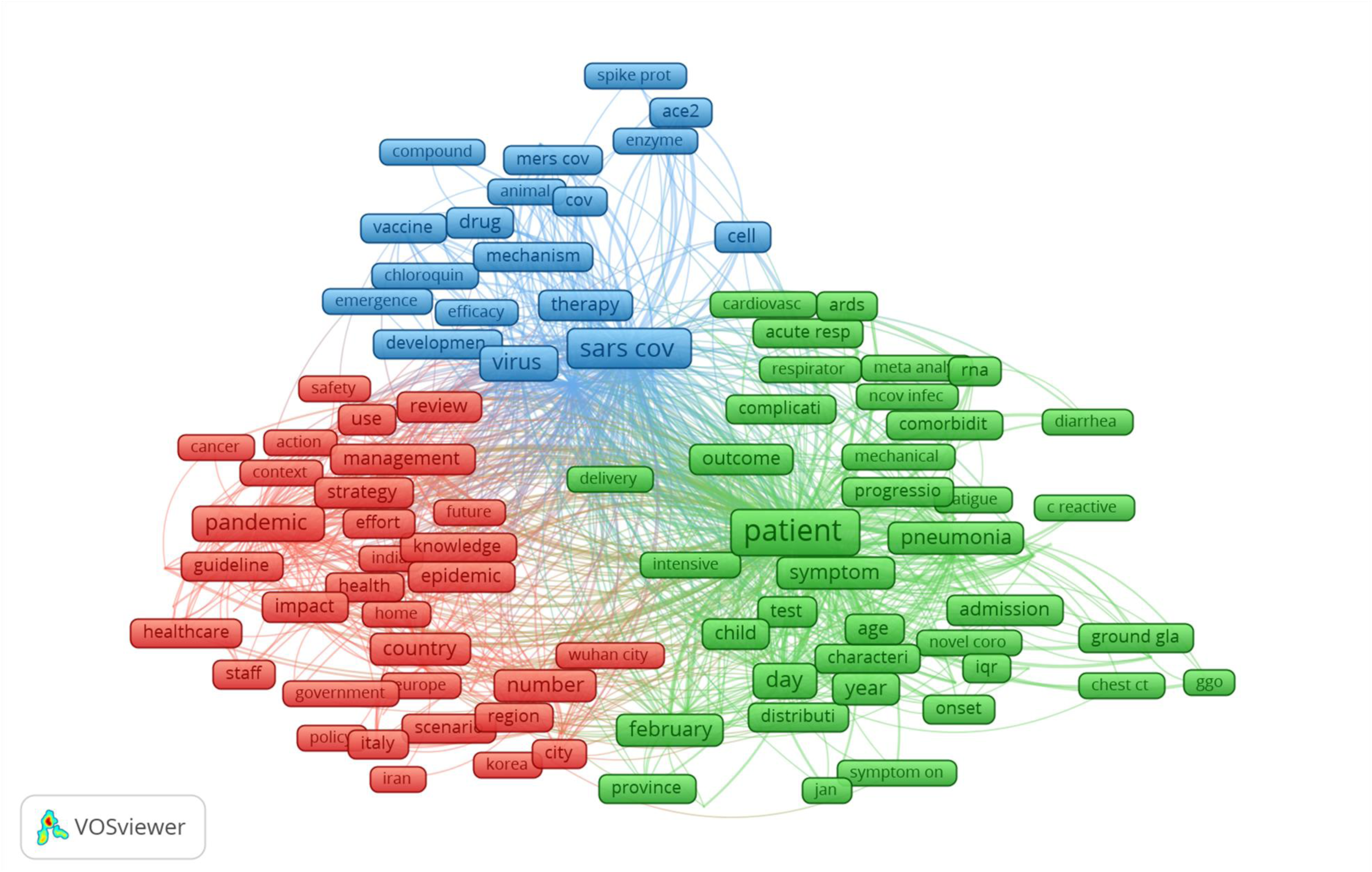
Co-occurrence analysis of the most frequent terms.

**Figure 5** shows the clustering of research areas in the WOS database. The research landscapes in this research field was the combination of several research areas. First cluster was Infectious diseases and Pharmacology. This cluster had a close connection with Surgery and Gastroenterology (second cluster). The third cluster related to treatment and diagnosis (such as Radioogy, Hematology, Virology, Psychiatry, Gerontology, or Metanolism). The other clusters in COVID-19 research areas, including 1) Critical car and Respiratory System (the fourth cluster); 2) Health care service and health policy (the fifth cluster); 3) Microbiology and Immunology (the sixth cluster); 4) Oncology and Experimental Research (the seven cluster); and 5) Biology.

**Figure 5.**
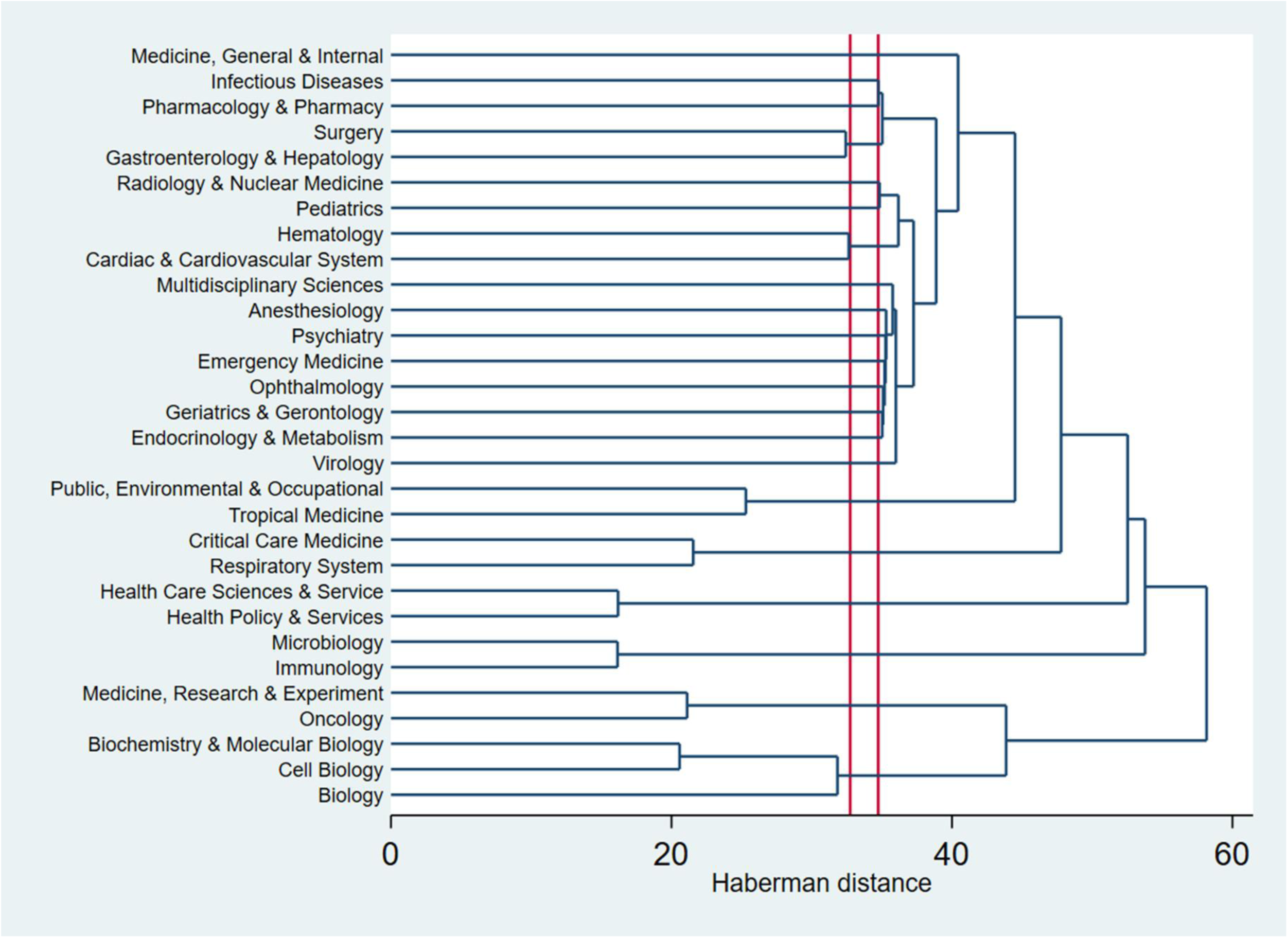
Dendrogram of research areas in WOS database.

The LDA results are presented in **Table 2**. Since the beginning of the pandemic, researchers have devoted special attention to the biology of SARS-CoV-2 (Topic 3 and 4) and made an enormous effort on various aspects of clinical investigation, such as diagnostic tests for detection of the virus, clinical examination of hospitalized patients and treatment for the disease (Topic 5, 7, 8, 9, 10, 11, and 15). Meanwhile, research on global and national responses to COVID-19 accounted for nearly a quarter of available literature volume (Topic 2, 12, and 13). Epidemiological characterization of COVID-19 and psychological disorders during the epidemic are also of great interest (Topic 1, 6, and 14).

**Table 2.** Fifteen topics about COVID-19 according to topic modeling

| Topic | Content | Top ten most frequent terms | n | % |
| --- | --- | --- | --- | --- |
| Topic 1 | Epidemiological reports on COVID-19 outbreaks in different countries | covid-19; cases; transmission; first; disease; coronavirus; march; countries; confirmed; health | 295 | 5.1 |
| Topic 2 | Global and international health security and responses in COVID-19 pandemic crisis | health; covid-19; pandemic; public; global; response; community; world; emergency; outbreak | 571 | 9.9 |
| Topic 3 | SARS-CoV-2 virus structure and molecular analysis | sars-cov-2; 2019-ncov; human; coronavirus; sars-cov; protein; virus; viral; spike; receptor | 231 | 4.0 |
| Topic 4 | Distinguishment between old and novel coronavirus: origin, pathology and pathogenesis | coronavirus; respiratory; novel; china; disease; sars-cov-2; severe; syndrome; acute; outbreak | 611 | 10.6 |
| Topic 5 | Radiographic detection of COVID-19 | covid-19; patients; symptoms; pneumonia; chest; clinical; disease; children; findings; imaging | 310 | 5.4 |
| Topic 6 | Psychological disorders in COVID-19 epidemic: epidemiological characteristics and interventions | covid-19; health; mental; during; outbreak; study; social; psychological; anxiety; media | 256 | 4.4 |
| Topic 7 | Clinical and laboratory examinations in hospitalized patients with COVID-19 | patients; covid-19; clinical; severe; study; group; cases; disease; results; wuhan | 232 | 4.0 |
| Topic 8 | Comorbidities in patients with COVID-19 | covid-19; patients; disease; respiratory; severe; acute; infection; syndrome; article; coronavirus | 474 | 8.2 |
| Topic 9 | Impacts of COVID-19 on pregnancy outcomes | covid-19; review; studies; evidence; women; pregnant; clinical; research; literature; results | 156 | 2.7 |
| Topic 10 | Diagnostic values of SARS-CoV-2 tests and improvement strategies | sars-cov-2; positive; covid-19; viral; testing; detection; rt-pcr; samples; results; negative | 234 | 4.1 |
| Topic 11 | Guidelines for emergency care and surgical management during COVID-19 pandemic | covid-19; pandemic; patients; during; management; cancer; hospital; recommendations; clinical; surgery | 669 | 11.6 |
| Topic 12 | Global logistics concerns in COVID-19 prevention, treatment and care | covid-19; protective; transmission; healthcare; workers; equipment; personal; during; infection; staff | 242 | 4.2 |
| Topic 13 | Medical education in COVID-19 pandemic | covid-19; pandemic; medical; education; medicine; during; response; lessons; students; nursing | 561 | 9.7 |
| Topic 14 | COVID-19 epidemiological modelling and forecasting | covid-19; cases; china; epidemic; number; outbreak; model; wuhan; measures; confirmed | 292 | 5.1 |
| Topic 15 | Treatment interventions against COVID-19 | covid-19; treatment; drugs; clinical; therapeutic; against; antiviral; sars-cov-2; therapy; effective | 311 | 5.4 |

The research focuses on countries with different income level and epidemic characteristics are shown in Table 3. High-income countries (HICs) were significantly more likely to report fewer studies focused on research in epidemiological characteristics and interventions of Psychological disorders in COVID-19 pandemic (Topic 6) compared with countries with other income levels. Low-middle income countries were found to publish less on diagnostic values of SARS-CoV-2 tests and improvement strategies (Topic 10). Treatment interventions for COVID-19 (Topic 15) attracted the interest of scientists among countries at all income levels, especially in HICs.

**Table 3.**
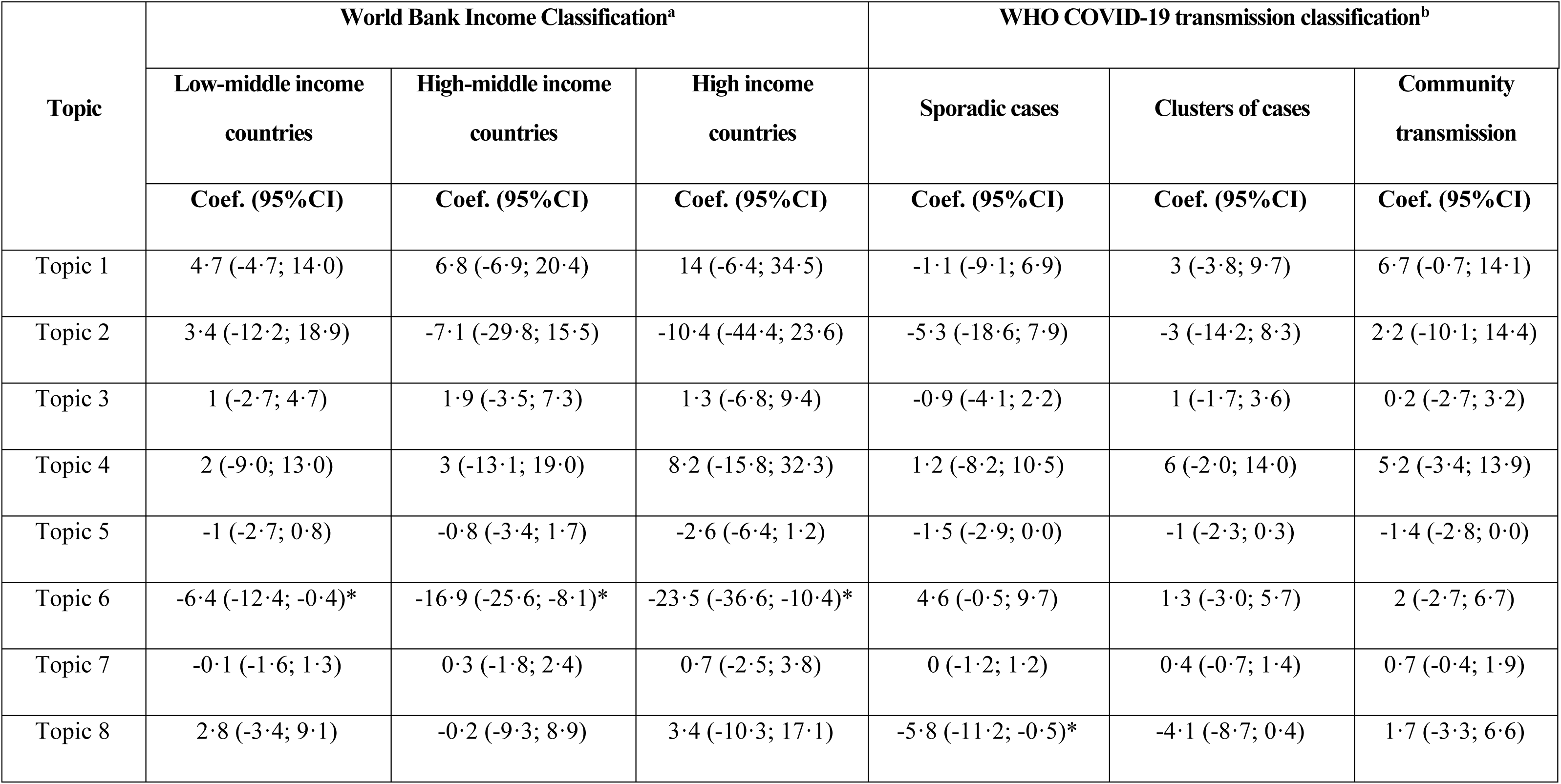
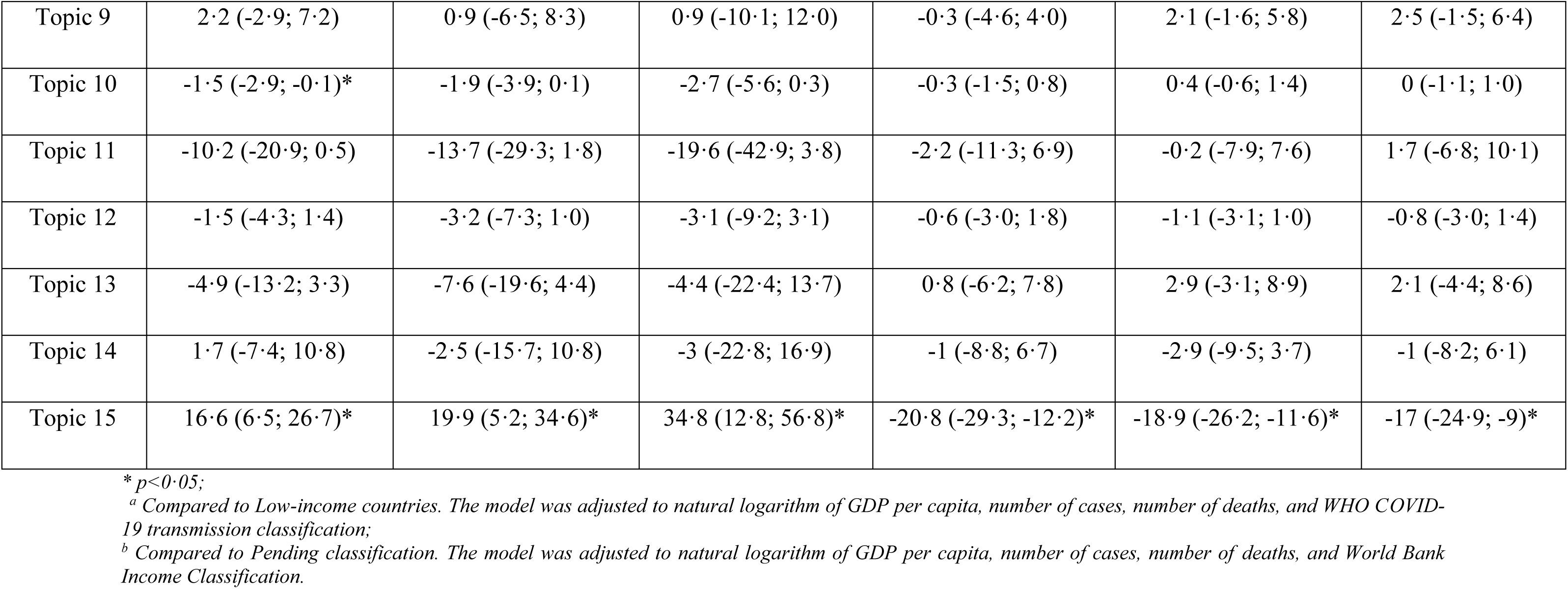
Regression models to identify the research trend among countries with different income level and epidemic characteristics

| Topic | World Bank Income Classification<sup>a</sup> |  |  | WHO COVID-19 transmission classification<sup>b</sup> |  |  |
| --- | --- | --- | --- | --- | --- | --- |
|  | Low-middle income countries | High-middle income countries | High income countries | Sporadic cases | Clusters of cases | Community transmission |
|  | Coef. (95%CI) | Coef. (95%CI) | Coef. (95%CI) | Coef. (95%CI) | Coef. (95%CI) | Coef. (95%CI) |
| Topic 1 | 4.7 (-4.7; 14.0) | 6.8 (-6.9; 20.4) | 14 (-6.4; 34.5) | -1.1 (-9.1; 6.9) | 3 (-3.8; 9.7) | 6.7 (-0.7; 14.1) |
| Topic 2 | 3.4 (-12.2; 18.9) | -7.1 (-29.8; 15.5) | -10.4 (-44.4; 23.6) | -5.3 (-18.6; 7.9) | -3 (-14.2; 8.3) | 2.2 (-10.1; 14.4) |
| Topic 3 | 1 (-2.7; 4.7) | 1.9 (-3.5; 7.3) | 1.3 (-6.8; 9.4) | -0.9 (-4.1; 2.2) | 1 (-1.7; 3.6) | 0.2 (-2.7; 3.2) |
| Topic 4 | 2 (-9.0; 13.0) | 3 (-13.1; 19.0) | 8.2 (-15.8; 32.3) | 1.2 (-8.2; 10.5) | 6 (-2.0; 14.0) | 5.2 (-3.4; 13.9) |
| Topic 5 | -1 (-2.7; 0.8) | -0.8 (-3.4; 1.7) | -2.6 (-6.4; 1.2) | -1.5 (-2.9; 0.0) | -1 (-2.3; 0.3) | -1.4 (-2.8; 0.0) |
| Topic 6 | -6.4 (-12.4; -0.4)* | -16.9 (-25.6; -8.1)* | -23.5 (-36.6; -10.4)* | 4.6 (-0.5; 9.7) | 1.3 (-3.0; 5.7) | 2 (-2.7; 6.7) |
| Topic 7 | -0.1 (-1.6; 1.3) | 0.3 (-1.8; 2.4) | 0.7 (-2.5; 3.8) | 0 (-1.2; 1.2) | 0.4 (-0.7; 1.4) | 0.7 (-0.4; 1.9) |
| Topic 8 | 2.8 (-3.4; 9.1) | -0.2 (-9.3; 8.9) | 3.4 (-10.3; 17.1) | -5.8 (-11.2; -0.5)* | -4.1 (-8.7; 0.4) | 1.7 (-3.3; 6.6) |
| Topic 9 | 2.2 (-2.9; 7.2) | 0.9 (-6.5; 8.3) | 0.9 (-10.1; 12.0) | -0.3 (-4.6; 4.0) | 2.1 (-1.6; 5.8) | 2.5 (-1.5; 6.4) |
| Topic 10 | -1.5 (-2.9; -0.1)* | -1.9 (-3.9; 0.1) | -2.7 (-5.6; 0.3) | -0.3 (-1.5; 0.8) | 0.4 (-0.6; 1.4) | 0 (-1.1; 1.0) |
| Topic 11 | -10.2 (-20.9; 0.5) | -13.7 (-29.3; 1.8) | -19.6 (-42.9; 3.8) | -2.2 (-11.3; 6.9) | -0.2 (-7.9; 7.6) | 1.7 (-6.8; 10.1) |
| Topic 12 | -1.5 (-4.3; 1.4) | -3.2 (-7.3; 1.0) | -3.1 (-9.2; 3.1) | -0.6 (-3.0; 1.8) | -1.1 (-3.1; 1.0) | -0.8 (-3.0; 1.4) |
| Topic 13 | -4.9 (-13.2; 3.3) | -7.6 (-19.6; 4.4) | -4.4 (-22.4; 13.7) | 0.8 (-6.2; 7.8) | 2.9 (-3.1; 8.9) | 2.1 (-4.4; 8.6) |
| Topic 14 | 1.7 (-7.4; 10.8) | -2.5 (-15.7; 10.8) | -3 (-22.8; 16.9) | -1 (-8.8; 6.7) | -2.9 (-9.5; 3.7) | -1 (-8.2; 6.1) |
| Topic 15 | 16.6 (6.5; 26.7)* | 19.9 (5.2; 34.6)* | 34.8 (12.8; 56.8)* | -20.8 (-29.3; -12.2)* | -18.9 (-26.2; -11.6)* | -17 (-24.9; -9)* |

\* $p < 0.05$ ;
<sup>a</sup> Compared to Low-income countries. The model was adjusted to natural logarithm of GDP per capita, number of cases, number of deaths, and WHO COVID-19 transmission classification;
<sup>b</sup> Compared to Pending classification. The model was adjusted to natural logarithm of GDP per capita, number of cases, number of deaths, and World Bank Income Classification.

Regarding transmission classification, comorbidities in patients with COVID-19 (Topic 8) were found to receive less attention among countries with sporadic cases in comparison with countries having “pending” transmission classification. Treatment interventions were significantly less likely to be the research focused in countries having sporadic cases, a cluster of cases, and community transmission compared with those with “pending” transmission classification.

## Discussion

By using the natural language processing approach with the Latent Dirichlet allocation, this study was able to capture the focus of COVID-19 related publications in different settings. This paper informed the rapid growth of research publications, the global variation in research productivity, and research interests. Moreover, the findings of this study suggest research gaps.

In this study, we found a greater number of publications regarding COVID-19 and SARS-CoV-2 in comparison with previous bibliometric studies.^21-23^ For example, Lou et al. used the Medline database and only found 183 publications through February 29, 2020.^22^ This disparity could be justified that our search was far more comprehensive than these studies by usingthree major databases, including the Medline, Scopus, and Web of Science. In addition, we included other document types such as letters, commentaries, or notes rather than concentrating only on original articles. As original papers require a long period for peer-reviewed,^30^ scientists tended to publish their ideas in those document types first for receiving rapid feedbacks from others,^31^ Therefore, we believed that our approach was appropriate given that these documents might partly reflect the research focus in each country.

The thematic maps of authors’ keywords and terms reveal that major research themes included virological and molecular analysis of the virus; clinical, laboratory and radiology examinations; and global and public health responses. Our findings are in line with a previous bibliometric study, which showed that virology, clinical characteristics, and epidemiology of COVID-19 were found to be the research focus with the highest volume of papers.^22^ Indeed, these research areas are essential components for controlling the pandemic. While understanding the biology of SARS-CoV-2 is critical for the development of effective and safe screening tests, drugs and vaccines. Investigations into clinical characteristics of COVID-19, along with results of laboratory and radiology, could inform a fundamental for appropriate patient management in the clinical settings. Research on public health responses could illustrate the effectiveness of different policies and strategies to mitigate the consequences of COVID-19 pandemic.^32-34^

The results of topic modeling offer more penetrating insights into the emerging research themes. Of all identified topics, clinical aspects, particularly guidelines for emergency care and surgical management during the COVID-19 pandemic (Topic 11), were most frequent. Along with the rapid increase in the number of confirmed cases, the heavy demand for health facilities and health workers, as well as the lack of effective treatment regimens, place a heavy burden and prevent the healthcare systems from operating efficiently. Without guidelines for prompt responses in emergency care, the burden caused by COVID-19 would go beyond the capacity of most health systems, especially for ICU care.^35^ In addition, a number of SARS-CoV-2 infections emerged from operations were reported in China, suggesting the risk of virus exposure despite strict hygienic requirements and aseptic techniques during the surgical process.^36^ Research for clinical guidelines, therefore, plays a critical role in mitigating the impact of COVID-19 on the healthcare system.

The origin and pathophysiology of the virus have attracted a great deal of attention since the beginning of the outbreak.^37-39^ The interest on this topic has continued to rise as the virus has gone beyond China, where the first infection was reported, and positive cases have been found in most countries and territories.^40^ On the other hand, the information that SARS-CoV-2 is a laboratory derived virus, albeit has been confirmed to be a false claim, gave rise to considerable controversy and also facilitated research on the nature of the virus ^41^. Another topic that should be mentioned is national public health responses and actions against COVID-19, especially at the beginning of the pandemic, when there was a wide difference in policies introduced by different governments. In particular, some countries have been advocating achieving herd immunity, whereas low- and middle-income countries (LMICs) have implemented strict actions, including quarantine, isolation, social distancing, and community containment as soon as the outbreak occurred ^42-45^. Although such measures have sdemonstrated their effectiveness, for optimal public health as well as economic outcomes, further investigation into their implementation within specific contextual factors should be prioritized.^46^ Moreover, continued medical training for healthcare workers ^47^ and preventive measures for the workforce,^48^ along with frequent transparent communication and educational interventions for the public is essential to strengthen the preventive capacity of each individual and thus, contribute to the global fight against COVID-19. Meanwhile, since COVID-19 has been reported to have no noticeable effect on pregnancy, research on COVID-19 among pregnant women received relatively slight interest.^49^

Regarding the research trend and level of interest of different country groups, it appeared that the share of publications regarding the topic about psychological health and related interventions was negatively associated with the income levels. This finding might imply that this topic might not be the priority of the countries, or in other words, developed nations even show less interest than the ones having lower-income.^50^ However, we do believe that most of the studies about this topic were cross-sectional surveys in the community, which were more affordable for low-income countries to perform compared to other topics. Nevertheless, COVID-19 caused a significantly psychiatric impact,^51^ and this impact maintained when the total number of COVID-19 cases continues to rise in a country.^52^ Developed nations are not immune from mental health issues and mental health services are often disrupted during COVID-19 pandemic.^53^ In terms of treatment interventions, although all countries are making an effort to develop an effective treatment regimen, high-income countries, with their vast financial resources, greater expertise, and infrastructure, demonstrated their bold attempt at this research area.^54,55^ Meanwhile, compared to low-income nations, we observed a lower share of SARS-CoV-2 test-related publications among low-middle income countries, suggesting that these countries prioritized to other research fields such as treatment interventions given the resource-constrained.^56^

Findings also suggested that research on comorbidities associated with COVID-19 is relatively underdeveloped in countries with sporadic cases, in contrast with the extensive understanding and research on the effects of comorbidities on COVID-19 of those countries with a high number of infections.^57,58^ On the other hand, treatment interventions were not prioritized, while the level of transmission increases. Although some high-income countries such as the United States, Canada or the United Kingdom were classified as “community transmission” and greatly contributed to the progress of finding treatment interventions, most of the nations in this categories were low-middle income countries (e.g. South American and African countries) and the governments tends to focus on preventive methods to prevent the pandemic from getting worse.^2^

This study has several implications. To begin with, while rapid transmission of COVID-19 has been triggering a strong need for the development of an effective vaccine, our results show minimalresearch on this research topic. In addition, since there have been anecdotalevidence that promising drugs for COVID-19, such as Lopinavir/ritonavir (LPV/RTV). Chloroquine (CQ) and hydroxychloroquine (H0), show no significant benefits to health outcomes of patients, hence,eveloping an effective and safe medication specific for the treatment of COVID-19 is of utmost importance ^59-61^. We also found a lack of research on the social stigma caused by COVID-19. Due to the rapid contagion of the virus, fear and anxiety about being infected can give rise to stigma and discrimination toward people, places, or things. For instance, people associated with the disease, such as being in the neighborhood of high risk or being civils of nations with a high rate of COVID-19, are often stigmatized.^62,63^ Stigma can also arise when a person is released from quarantine, even though they have been confirmed to be negative and are no longera risk. Although there have been several published guidelines for reducing social stigma related to COVID-19, further investigation into the detrimental effects of social stigma and development of interventions for this problem should be carefully considered.^62,63^

To our knowledge, this is the first analysis using text mining and text modeling to investigate the topics of the worldwide COVID-19 publications. However, some limitations should be notedd. The restriction of the search strategy to the English language might not reflect globalized practices and the research priority of a country, such as articles in the local language. Analyses of keywords, titles, and abstracts may not fully reflect the content of articles. However, with the combination of three large datasets and various techniques of text mining, this study is useful for an overview of the research direction.

## Conclusions

This study showed that COVID-19 related publications were primarily contributed by major research hubs such as the United States, China, and European countries. Global researchers were currently focusing on clinical management, viral pathogenesis, and public health responses in combating against COVID-19. We alsofound country variation in research priorities. Findings suggest the need for global research collaboration among high- and low/middle-income countries in the different stages of prevention and control the pandemic.

## Data Availability

Data is available from corresponding author upon reasonable request

## Authors’ contributions

Bach Xuan Tran: conceptualization, revision of manuscript; Giang Hai Ha: conceptualization, literature search, and writing; Hoang Long Nguyen: data analysis, literature search and writing; Giang Thu Vu: literature search, data analysis and writing; Hai Thanh Phan: literature search and writing; Huong Thi Le: revision of manuscript; Carl A. Latkin: revision of manuscript; Roger C.M. Ho: revision of manuscript; Cyrus S.H. Ho: revision of manuscript.

